# Utility of Polygenic Risk Scores in Families with Exceptional Longevity

**DOI:** 10.1101/2025.07.21.25331660

**Authors:** Laura Xicota, Rong Cheng, Sandra Barral, Lawrence S Honig, Nicole Schupf, Yian Gu, Stacy Andersen, Stephanie Cosentino, Joseph Zmuda, Thomas Perls, Michael Province, Joseph H Lee, Long Life Family Study (LLFS)

## Abstract

**Introduction:** Polygenic risk scores (PRS) have been used to assess an individual’s risk for various diseases, including Alzheimer’s disease (AD). This study applied the PRS approach to a cohort of families ascertained for healthy aging, that have shown a reduced risk of AD. Using the SNPs identified as significantly associated with AD in the study by Kunkle and colleagues, we examined the utility of PRS for predicting AD risk in a cohort ascertained for familial healthy aging.

**Methods:** We restricted the study to US LLFS study participants who have been evaluated for AD and have available whole genome sequencing (WGS) data. AD diagnosis was based on consensus diagnosis, and for those without consensus diagnosis, we used the algorithm based on standardized memory scores. PRS were calculated using a published weighted formula. To further examine the predictability of PRS, we assessed the relationship between PRS and AD biomarkers, including Aβ_42_, Aβ_40_, NfL, and GFAP. Mixed effects models were used to adjust for confounders as well as relatedness among family members. Given the age at onset of common late onset AD, the present study included those who were at least 65 years of age.

**Results:** We observed that PRS had limited predictive power for AD in this healthy aging cohort. Yet, allele frequencies for the SNPs used in PRS estimation differed between the two studies in a small number (9.7%) of SNPs, suggesting that the lack of effect of the PRS is likely to be due to the small number of AD associated SNPs (12.3% and 16.1%). Subsequent analysis observed no significant association between PRS and biomarkers. This was explained by the low number of SNPs significantly associated with each of the biomarkers.

**Conclusions:** This study highlights the importance of ascertainment of study population in interpreting PRS. In LLFS, a population at reduced risk of AD, PRS based on genetic variants identified from the general population may be inadequate to explain the variability in AD risk. Our results suggest that genetic risk variants, the basis of PRS, may need to be adjusted according to the study population of interest.

## INTRODUCTION

To date, multiple genome wide association studies (GWAS) of Alzheimer’s disease (AD) have identified a collection of candidate risk genes and variants within^1,2^. Studies have advocated the use of polygenic risk score (PRS) to assess an individual’s genetic susceptibility to AD and related traits, serving as the basis for personalized medicine to provide a global value of risk of disease such as AD and related phenotypes^1-10^. Excluding autosomal dominant forms of AD, most of the late onset AD (LOAD) are likely to be caused by multiple variants with small effects, and PRSs incorporate this feature of genetic complexity. Some have discussed concerns related to the complexity including among others, differences in populations’ s genetic background^11-13^. In the present study, we evaluate the utility of PRS in a cohort ascertained for familial exceptional healthy aging, namely the Long-Life Family Study (LLFS).

The LLFS cohort is a cohort of familial extreme longevity with reduced prevalence of AD^14^. The family members of LLFS have been shown to be resilient to major chronic diseases, including AD and other dementias^15^. Some studies have compared the genetics of these centenarian populations and found them to have reduced genetic risk factors for AD,^16-18^ which would explain their lower risk of dementia. At the same time, these genetic risk factors, in the form of PRS, could not explain AD cases in this protected population^16^.

In this study, we first evaluated the usefulness of PRS in the LLFS cohort, exploring the specific genetic characteristics of this cohort including allele frequencies and genetic associations with AD. To examine further the biological influence of PRS, we then examined both PRS and genetic risk factors and their relations to plasma biomarkers of AD, including Aβ_40_, Aβ_42_, Aβ_42_/Aβ_40_, NfL, and GFAP. Assessing genetic risk factors in populations like LLFS can highlight specific biological pathways that confer risk or protection against AD in centenarians.

## METHODS

### Study Participants

We studied US non-Hispanic White participants from LLFS who had whole genome sequence (WGS) data available and a diagnosis of Alzheimer’s disease diagnosis, totaling 3,408 family members. Participants from the Danish site were not included in the analysis. In addition, we also performed an age-restricted analysis with participants who were 65 years of age or older to exclude participants who were not at risk of late onset AD. The final sample for analysis yielded a total of 2,482 participants. Characteristics of the LLFS cohort have been described elsewhere^14,19^.

### Cognitive measures

Alzheimer’s disease diagnosis was obtained via consensus diagnosis, as previously described^20^. For individuals without consensus diagnosis, probable AD diagnosis was determined using a diagnostic algorithm^14^ and selecting those with anomalous standardized memory scores. The rest of the participants were considered to be non-AD.

### Whole genome sequencing

Whole genome sequencing processing was performed^20,21^ by the McDonnell Genome Institute (MGI) at Washington University in St. Louis via 150bp reads by Illumina Sequencers. Subsequently, MGI performed sequence alignment to build GRCh38 with BWA-MEM, marking duplicates with Picard, base quality score recalibration with Genome Analysis Toolkit (GATK), and lossless conversion to CRAM format with SAM tools. Variant calling and quality control metrics were performed at the LLFS Data Coordinating Center, Division of Statistical Genomics at Washington University in St. Louis, using GATK4.1.0.0.

### Biomarkers

Blood was obtained as per LLFS procedures during home visits to participants, via venipuncture, drawn in Potassium EDTA tubes. Tubes were shipped at 4°C overnight to the central sample repository, where blood was centrifuged and plasma divided into 250-500 uL aliquots in polypropylene cryotubes and stored at -70°C. The Quanterix HD-X platform was used with the following kits: Simoa Neurology 4-plex E (N4PE Catalog No. 103670) for Aβ40, Aβ42, NfL, and GFAP. All samples were measured in duplicate, and any samples with coefficients of variation (COV) >10% were re-measured. Overall COV averaged 4.3%, 2.6%, 4.4%, and 4. 9%, for Aβ42, Aβ40, NfL, and GFAP. Biomarker and AD diagnosis data were available for 2,548 participants, where 1,408 of those were older than 65. The Aβ_42_/Aβ_40_ ratio was computed using concentration values of the amyloid peptides and ratios above 0.3 were considered missing.

### PRS calculation

Polygenic risk score (PRS) was calculated using a weighted method ^10^, with the single nucleotide polymorphisms (SNPs) identified by Kunkle et al that were deemed to be genome-wide significant^2^. To account for family structure, the analysis included genetic relatdenss matrix (GRM) as detailed below. Of the 33 SNPs presented on the paper, 31 passed our quality control measures and were included in the analysis. PRS was calculated for those participants who had information for all the SNPs, that is, 2,320 participants were included in the AD-risk main analyses and 1,627 (65.6%) in the age-restricted sub-analyses (65.2%). To further assess the effects of PRS, we first compared the AD risk associated with the top 45 percentile in PRS against the bottom 45%, treating the middle 10% as unknown to minimize the effects of misclassification. We then repeated the analysis using the 10 percentiles as the threshold cutpoint where we compared the top 10% against the bottom 85%, treating the remaining 5% as unknown.

### Allele frequency

We estimated allele frequencies by restricting the samples to unrelated founders as implemented in PLINK 1.9.^22^ Comparisons between allele frequencies were performed using χ^2^ or Fisher’s exact test.

### Statistical analysis

The genetic relationship matrix (GRM) was generated using GEMMA^23^ to account for relatedness among family members. Subsequently, a linear mixed effects model was used to examine the association between AD and PRS or SNP. This modeling was implemented in GMMAT^24^. All biomarkers, except for the Aβ_42_/Aβ_40_ ratio, were transformed using log base-10 prior to performing regression models to fit required normality. Multivariable logistic regression analyses were adjusted for sex, age, education (<12 years vs ≥12), *APOE* ε4 status (carrier of at least one allele vs non-carrier), the first three genomic principal components (PC1 to PC3), and GRM. Multiple linear regression models were adjusted for sex, age, *APOE* ε4 status, body mass index (BMI), creatinine concentrations, PC1 to PC3, and GRM.

## RESULTS

### Demographic description

Compared with cognitively healthy participants, those affected with AD were significantly older (p<10^−4^) and less likely to have education beyond 12 years (p<10^−4^) (Table 1). No significant differences were observed in the distribution of sex or *APOE* allele frequency between AD cases and unaffecteds. The AD prevalence in the LLFS participants was 17% for the whole cohort and 23% for those older than 65 years of age. The majority (75%) of those who were affected were older than 87 years.

**Table 1:** Demographic and APOE characteristics of the study participants in LLFS.

|  |  | Total* |  |  | ≥65 years of age |  |  |
| --- | --- | --- | --- | --- | --- | --- | --- |
|  |  | AD<br>(N=567) | Non-AD<br>(N=2,841) |  | AD<br>(N=559) | Non-AD<br>(N=1,923) |  |
| Age | Mean±SD | 90.2±8.3 | 72.8±13.6 | **p<0.0001** | 90.8±7.6 | 79.6±10.8 | **p<0.0001** |
|  | Range | 47-108.8 | 24.9-110.5 |  | 65.4-108.8 | 65-110.5 |  |
| Sex | Females | 297 (15.8%) | 1,582<br>(84.2%) | p=0.15 | 293 (22.3%) | 1,024<br>(77.7%) | p=0.73 |
|  | Males | 270 (17.7%) | 1,259<br>(82.3%) |  | 266 (22.8%) | 899 (77.1%) |  |
| Education** | <12 years | 326 (29.1%) | 793 (70.9%) | **p<0.0001** | 322 (33.4%) | 643 (66.6%) | **p<0.0001** |
|  | ≥12 years | 239 (10.5<br>%) | 2,043<br>(89.5%) |  | 235 (15.6%) | 1,276<br>(84.4%) |  |
| APOE2 frequency |  | 7.7% | 7.7% | p=0.34 | 7.9% | 9.2% | p=0.33 |
| APOE4 frequency |  | 8.6% | 8.6% | p=0.84 | 8.3% | 8.4% | p=0.88 |

\*The table presents both total population and age-restricted to those 65 and older. Significant differences are presented in bold.
\*\*Information on the level of education was not available for all participants.

### PRS was associated with AD status in the LLFS population

Prior to examining the relative usefulness of PRS in LLFS to predict AD, we performed a series of association analysis using one risk factor at a time, including age, sex, education, *APOE*-ε4 carrier status, while considering PC1 to PC3 and GRM, to assess how much PRS adds to known risk factors for AD.

As shown in Table 2, age (OR=1.12, p<10^−4^) was a strong risk factor for AD, as expected, while a high level of education, defined as ≥12 years of schooling, was a significant protective factor (OR=0.28, p<10^−4^) against AD. It is further noted that the OR for age was slightly higher than that for PRS (OR=1.04, p=0.073), suggesting it has a greater discriminating power (Table 2). However, PRS with the four covariates had a comparable but nominally significant OR (OR=1.07, p=0.012), however the two variables together did not outperform age alone. We then created PRS categories to determine whether there exists a subset of high-risk individuals represented by high PRS beyond a certain threshold. To this end, we compared the top 45 percentile PRS as a high-risk group against the rest. This analysis yielded an OR of 1.36 of having AD (p=0.034). When we elevated the threshold to the top 10 percentile and compared them to the rest, those with the top 10 percentile PRS had a 1.6-fold higher risk of developing AD (OR=1.60, p=0.031). The observed threshold effect yielded similar ORs whether we restricted the analysis to the older group (Table 2).

**Table 2:** PRS associated Alzheimer’s disease risk in all family members vs. those who are 65 years of age or older. Significant values are presented in bold. Results are univariate analyses unless otherwise indicated.

|  | LLFS - All |  |  | LLFS ≥65 years |  |  |
| --- | --- | --- | --- | --- | --- | --- |
|  | Beta | OR | p | Beta | OR | P |
| ***Known risk factors for AD*** |  |  |  |  |  |  |
| Age | **0.113** | **1.12** | **<0.0001** | **0.109** | **1.12** | **<0.0001** |
| Sex | -0.126 | 0.88 | 0.174 | -0.028 | 0.97 | 0.773 |
| Education (<12 vs ≥12 years) | **-1.278** | **0.28** | **<0.0001** | **-1.000** | **0.37** | **<0.0001** |
| APOE ε4 (carrier vs non-carrier) | 0.015 | 1.02 | 0.906 | 0.052 | 1.05 | 0.697 |
| ***PRS + 4 Known Risk Factors*** |  |  |  |  |  |  |
| PRS | <u>0.038</u> | <u>1.04</u> | <u>0.073</u> | **0.052** | **1.05** | **0.020** |
| PRS with the above 4 covariates | **0.065** | **1.07** | **0.012** | **0.066** | **1.07** | **0.011** |
| PRS Top 45 | 0.163 | 1.18 | 0.179 | 0.207 | 1.23 | 0.103 |
| PRS Top 45 with covariates | **0.305** | **1.36** | **0.034** | **0.310** | **1.36** | **0.032** |
| PRS Top 10 | <u>0.334</u> | <u>1.40</u> | <u>0.060</u> | **0.434** | **1.54** | **0.022** |
| PRS Top 10 with covariates | **0.469** | **1.60** | **0.031** | **0.467** | **1.60** | **0.034** |

\*The middle 10% was considered missing to minimize misclassification.
\*\* Top 10% vs. bottom 85%. The remaining 5% was considered missing.

### Allele frequencies in LLFS differed only in a small number of SNPs

Prior to estimating PRS for AD in LLFS, we compared differences in reported minor allele frequencies (MAFs) for the informative SNPs generated from the study by Kunkle *et al*.^*2*^, against the LLFS cohort to determine whether the healthy aging cohort had lower allele frequencies of risk alleles. As shown in Table 3, of the 31 SNPs present in LLFS, three SNPs (9.7%) showed significantly different MAF between Kunkle et al and LLFS: rs71618613 in *SUCLG2P4*, rs2632516 in *MIR142/TSPOAP1-AS1*, and rs6024870 in *CASS4*. For these three SNPs, the minor allele was protective in Kunkle et al, while LLFS cohort exhibited was higher values. Additional six SNPs approached, but did not reach, statistical significance in LLFS (0.05<p-value<0.1), rs35868327 in FST, rs114812713 in OARD1, rs12539172 in NYAP1, rs4735340 in *NDUFAF6*, rs7933202 in *MS4A2*, and rs138190086 in *ACE* (See Table 3), for approximately one-fourth of the available SNPs. Of these, while rs114812713 in *OARD1* and rs138190086 in *ACE* were putative, the remaining four were protective (OR<1).

**Table 3:**
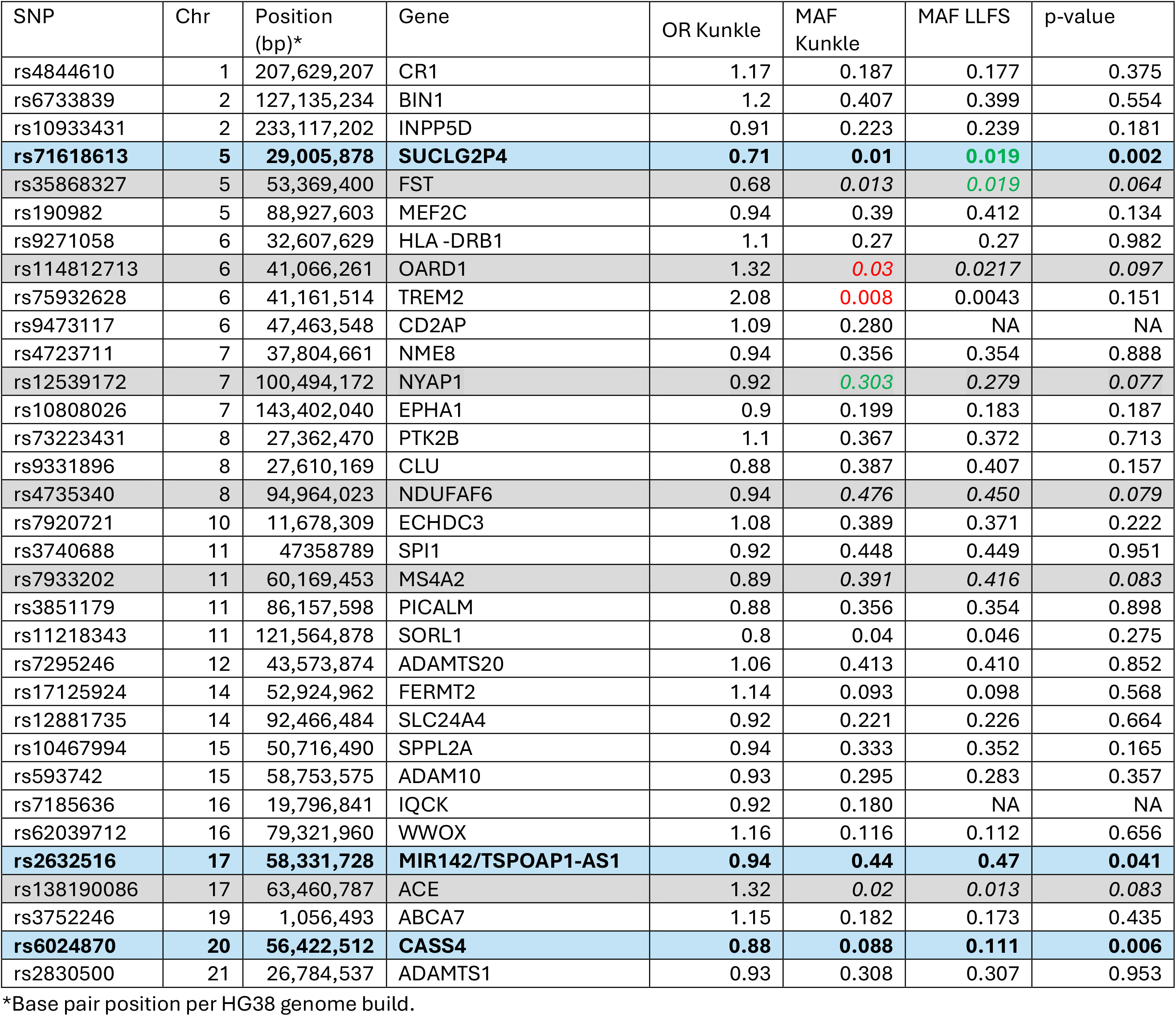
Differences in minor allele frequency Kunkle 2019 and the founder individuals in LLFS.

### Effect sizes were weaker in LLFS

Since PRS is a sum of sum of the product of each SNP’s genotype dosage and its corresponding effect size, we then compared the effect size for each SNP for LLFS against Kunkle et al. to determine whether the effect sizes of the risk variants in LLFS were weaker than that the reported ones. For risk variants, only two variants were significantly associated with AD at a larger OR in LLFS than in Kunkle et al. (Table 4). However, both were rare SNPs (MAF_TREM_=0.008, MAF_FERMT2_=0.093), suggesting their impact would be limited. The rest of the risk variants had lower OR_LLFS_ than OR_Kunkle_ with 59% of those having OR_LLFS_ below 1 (Table 5). In the case of protective variants, two variants were significantly associated with lower risk of AD in LLFS, with an additional one being significant in the older group (Table 4). In this case, two of the variants had high allele frequencies (MAF_NME8_=0.354, MAF_CLU_=0.407). The rest of the protective variants had OR_LLFS_ that were closer to 1, with 42% being above 1 (Table 5).

**Table 4:**
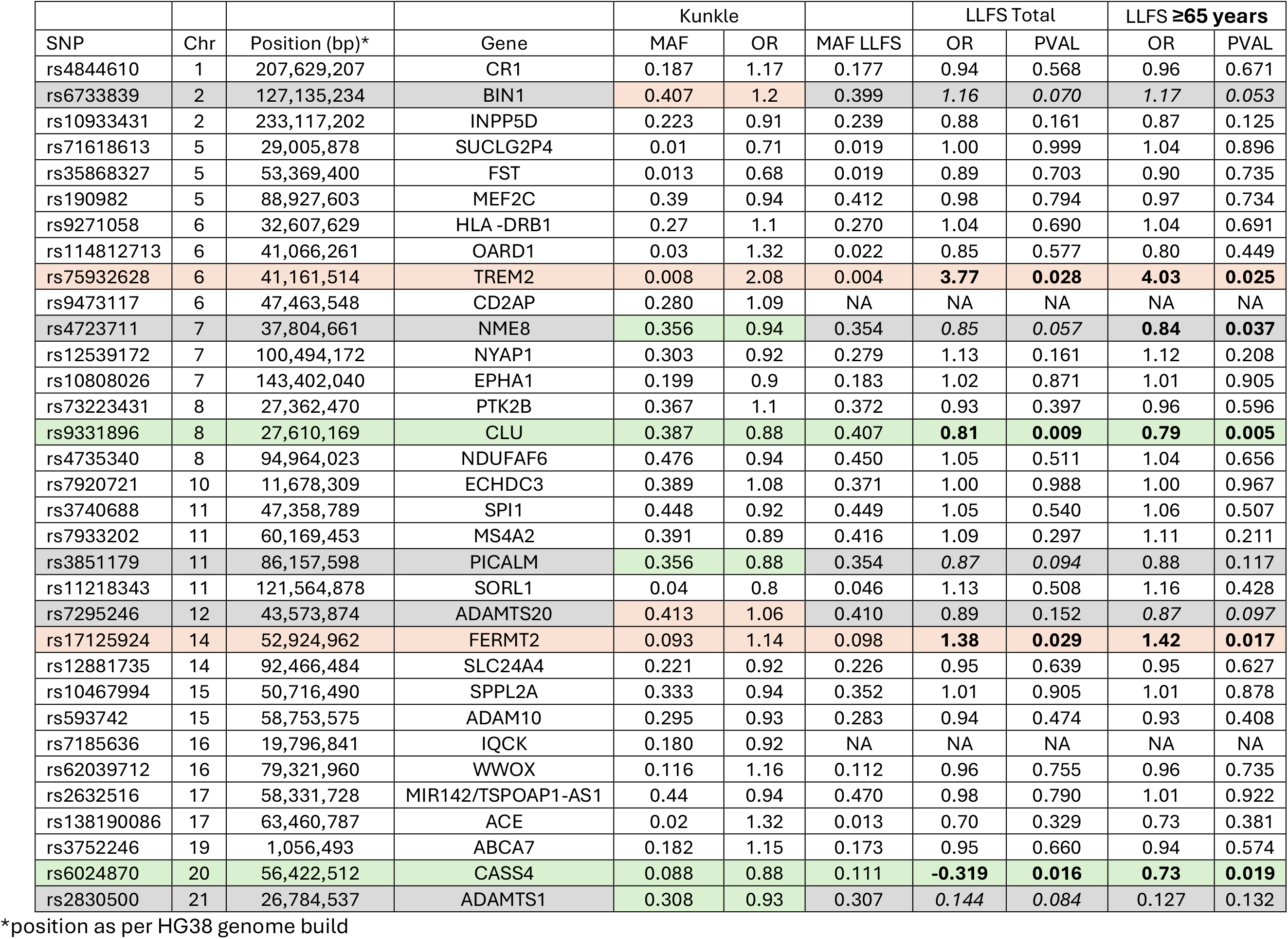
Comparison of effect sizes of the PRS SNPs and their association with AD in LLFS and Kunkle et colleagues.

**Table 5:**
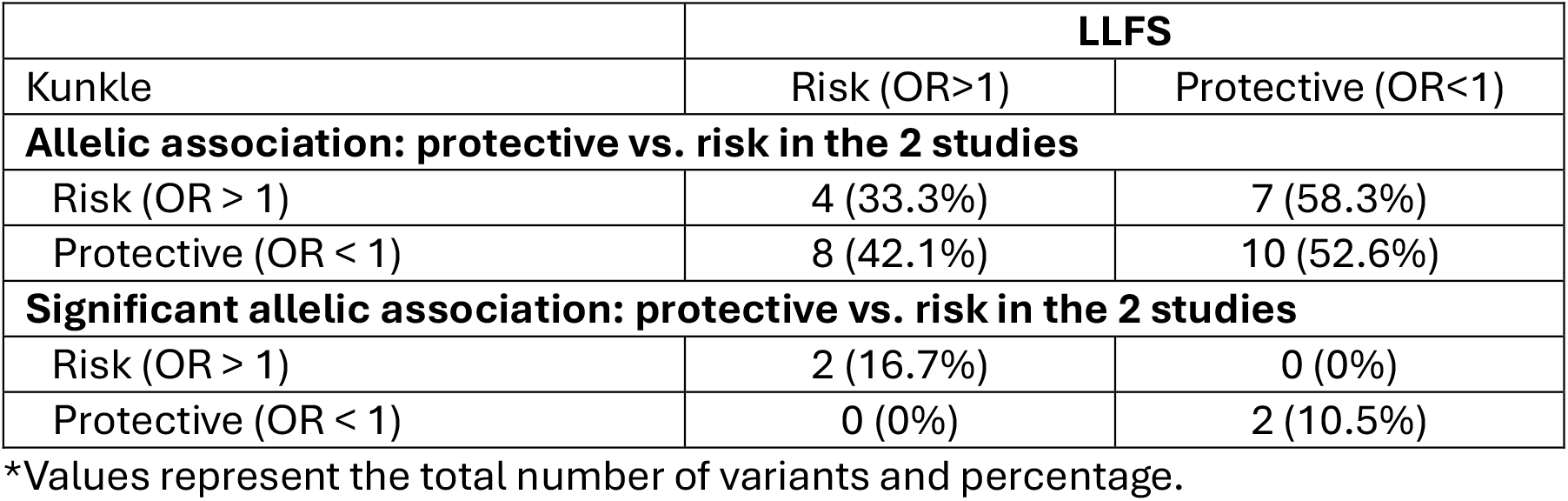
Comparison of SNP variants in LLFS and Kunkle et al: A summary of concordant vs. discordant effects*.

### PRS was not associated with biomarker endophenotypes

To further examine whether PRS has any biological impact on the phenotype, we evaluated the relations between PRS and plasma AD biomarkers (Supplementary Table 1). We did not observe any significant associations between PRS and AD biomarkers. We did observe marginal significance in association when comparing biomarker concentrations of those in the top 10% of PRS scores against the rest.

### AD risk variants were not associated with plasma biomarkers

To examine the lack of association between PRS and AD biomarkers, we performed single SNP association for each of the five available AD biomarkers. Of those, only five had significant associations with any of the biomarkers in both the primary analysis and the age-restricted (Table 7 and Supplement Table 2). None of the variants were significantly associated with AD in LLFS and only one had significant associations with more than one biomarker (*NYAP1* rs12539172 with higher concentrations of Aβ_40_ and Aβ_42_). This would explain the reasons for PRS not being associated with concentrations of AD biomarker.

## DISCUSSION

The present study showed that the PRS, derived from SNPs previously reported in a meta-analysis of the general population, was weakly associated with AD status. Moreover, PRS exhibited limited utility in predicting AD dementia risk in a familial healthy aging cohort. The lack of predictive power of PRS in LLFS was unlikely to be due to a lower allele frequency of risk alleles; rather, the number of AD associated SNPs in LLFS were significantly reduced. This suggests that some of these variants may have had functionally weak and unimpactful effects on LLFS participants harboring healthy aging traits. This could explain the observed lack of predictability of the PRS, and the reduced risk. Our findings were supported by the general lack of associations of both PRS and SNPs with plasma AD biomarkers.

Our earlier study on the LLFS revealed that family members are exceptionally healthy on multiple domains^19^, having a lower prevalence or delayed onset of certain diseases, including coronary heart disease,^25^ diabetes, depression, prostate cancer, heart failure, and chronic kidney disease^26^. Previous papers using a different AD classification have described a lower prevalence of AD in LLFS^14,26^. We earlier showed that a delayed onset of cognitive symptoms in individuals from exceptionally long-lived families when compared to their spouses^14^. Even though the prevalence of AD in LLFS was 23% in those older than 65 years of age, the majority (75%) of those who were affected were older than 87 years, supporting the notion that it is difficult to escape AD if you survive long enough.

To better understand the weak effect of PRS in our population, we first analyzed whether this population had different allele frequencies for the described SNPs. When the previously reported frequencies were compared with those in LLFS based on founder individuals (genetically independent samples), only three SNPs (10%) showed significantly difference, i.e., switch allele frequencies tended to be higher for LLFS. In all three cases, the minor allele was protective. These differences are not enough to explain the weak effect of PRS on our population. One particular study found associations with AD risk and protective variants and longevity; in some cases, the risk variants were associated with increased longevity^27^. The same group describes different allele frequencies of certain alleles list in centenarians as part of the Bellenguez et al. study ^17^, which we did not observe for those same SNPs (data not shown). One of the variants in which we observe increased allele frequency for the protective allele was rs6024870 in *CASS4*, previously reported as associated with increased longevity^27^, which would explain the higher allele frequency in our population.

Because the allele frequencies for PRS did not differ greatly between Kunkle et al. study and LLFS, we examined whether those SNPs were associated with AD in our population. In LLFS the percentage of SNPs associated with AD risk was low (12% for the principal analysis and 16% for the age-restricted one), which would explain the low predictability of the PRS. In all cases, the significant associations were in the same direction as what had been reported in Kunkle et al, with the protective effect described above. To our knowledge, no other studies have examined the association between AD risk SNPs and AD in healthy aging individuals.

When examining the associations of PRS with AD biomarkers, we did not observe any significant association. In addition, individual SNP associations provided more insight into the lack of associations with a limited number of SNPs being associated with any of the biomarkers. For the principal analysis, most of the significant SNPs (three) were associated with the Aβ_42/40_ ratio.

Although the lack of association observed in our population could be due to the fact that these were not biomarker-specific SNPs, other studies have shown associations between the same SNPs with AD plasma biomarkers. In one study that analyzed the associations between a modified version of the same SNP list r and plasma biomarkers, the authors described genetic associations between all biomarkers and a larger number of SNPs.^28^ One common association that we observed with this study was rs2830500 in *ADAMTS1* with Aβ_42/40_ ratio, where they presented only a trend towards significance. The rest of their significant associations failed to reach significance in the LLFS cohort.

This study highlights the impact of study participants ascertainment can influence PRS performance across different cohorts. Such is the case for the lack of association of *APOE-*ε4 with Alzheimer’s disease in this LLFS. The frequency of *APOE* -ε4 in the LLFS cohort is significant lower compared to Non-Hispanic White population (9% vs. ∼14%)^29^. This lower ε4 allele frequency could be explained by the detrimental effect that it has on longevity^30,31^. In addition, a similar lack of association of the ε4 allele with AD in centenarians has been previously described^32^. On the other hand, *APOE*-ε2 has been associated with extended longevity^33^ and lower risk of AD^34^ in LLFS, although slightly higher, the allele frequency does not differ as much from what has been published for several European populations^35^ (between 5 to 8% compared to 8% in LLFS).

Our work contributes to expanding previous studies examining PRS in centenarians such as LLFS cohort). Prior centenarian studies had described lower PRS scores for AD risk compared to the control population or AD patients^16-18^. However, the PRS was not associated with AD status in those who were exceptionally long-lived^16^. A recent study showed that centenarians had genetic protection against AD through a Polygenic Protective Score, which was also associated with decreased mortality^36^. In our study, we found an association between PRS and AD status, with an OR of 1.07 (p=0.012) per unit increment of PRS. This OR shows a relatively low effect on the risk of the PRS on AD in LLFS. In our population, the main predictor of AD status was age. In addition to performing the PRS based on the list of genes from Kunkle et al.,^2^ we performed the same set of analyses using the list of genes from a later GWAS by Bellenguez et al.^1^ (data not shown). As with PRS based SNPs from Kunkle et al. study, the PRS_Bellenguez_ had weaker associations and failed to predict AD status in LLFS. In the present study, we focused on the results from the Kunkle list as the AD-cases are confirmed AD rather than proxy. In studies in the general population, PRS scores have been shown to be associated with mild cognitive impairment (MCI) status^2,5^, AD status^2-4^, speed of conversion^6^, brain β-amyloid load^8,10^, and incident dementia^5,9^, as well as being capable of predicting conversion from MCI to AD^7^. Our findings suggest that when compared to general population, the LLFS cohort may harbor unique genetic variants contributing to AD risk.

We have examined a unique population of families with exceptional longevity, an ideal group in which to analyze genetic risk factors for AD. However, a limitation of our study is that we were unable to classify consensus diagnosis for participants without functional data. Additionally, certain biomarker data were unavailable at the time of analysis, including tau phosphorylation, CSF biomarkers or neuroimaging. This is in part due to the fact that participants are generally seen in a home setting, which did not allow for lumbar punctures or neuroimaging data collection.

The present study illustrates how ascertainment can have a significant impact on the utility of PRS in predicting AD. Specifically, we showed that PRS from the general population was not an adequate predictive tool in LLFS. This lack of impact on AD risk also translated into lack of association with biomarkers and therefore the underlying biology of the disease. In addition to the possible explanations mentioned above to explain the limited predictability in this cohort, several other possibilities can be considered such as the effect size of known variants may be altered due to having additional genetic modifiers may influence the phenotype of interest, resulting in weaker genotype-phenotype relations. Additional research will be needed to uncover the genetic variants that are associated with AD and its biomarkers in a healthy aging population, and to develop a cohort specific PRS.

## Data Availability

All data is being uploaded to the ELITE portal and will be available there. If there is immediate need for the data, contact the authors.

## ACKNOWLEDGMENTS

We are grateful to family members who are participating in the Long-Life Family Study and allowing us to study how they have achieved their healthy longevity. This work was funded by the National Institute on Aging/National Institutes of Health (grant numbers: U01-AG023746, U01-AG023712, U01-AG023749, U01-AG023755, U01-AG023744, and U19 AG063893)

## CONFLICT OF INTEREST

LSH is a consultant for Bioarctic, Biogen, Corium, Eisai, Genentech/Roche, Eli Lilly, Medscape, and New Amsterdam. LSH receives research funding from Acumen, Alector, Alnylam, Biogen, Bristol-Myers Squibb, Cervomed/EIP, Cognition, Eisai, Ferrer, GemVax, Genentech/Roche, GSK, Janssen/Johnson & Johnson, Eli Lilly, Transposon, UCB, Vaccinex, NIH/NIA, NIH/NINDS, and NYS.

## Notes

### Author Declarations

IRB at Washington University in St. Louis gave ethical approval for this work. IRB ID #201904204=1025.

## REFERENCES

1. Bellenguez C, Küçükali F, Jansen IE, et al. New insights into the genetic etiology of Alzheimer’s disease and related dementias. Nature Genetics 2022; 54(4): 412–36.

2. Kunkle BW, Grenier-Boley B, Sims R, et al. Genetic meta-analysis of diagnosed Alzheimer’s disease identifies new risk loci and implicates Aβ, tau, immunity and lipid processing. Nature Genetics 2019; 51(3): 414–30.

3. Altmann A, Scelsi MA, Shoai M, et al. A comprehensive analysis of methods for assessing polygenic burden on Alzheimer’s disease pathology and risk beyond APOE. Brain communications 2020; 2(1): fcz047.

4. Sleegers K, Bettens K, De Roeck A, et al. A 22-single nucleotide polymorphism Alzheimer’s disease risk score correlates with family history, onset age, and cerebrospinal fluid Aβ42. Alzheimer’s & dementia : the journal of the Alzheimer’s Association 2015; 11(12): 1452–60.

5. Adams HHH, de Bruijn RFAG, Hofman A, et al. Genetic risk of neurodegenerative diseases is associated with mild cognitive impairment and conversion to dementia. Alzheimer’s & Dementia 2015; 11(11): 1277–85.

6. Rodríguez-Rodríguez E, Sánchez-Juan P, Vázquez-Higuera JL, et al. Genetic risk score predicting accelerated progression from mild cognitive impairment to Alzheimer’s disease. Journal of neural transmission (Vienna, Austria : 1996) 2013; 120(5): 807–12.

7. Chaudhury S, Brookes KJ, Patel T, et al. Alzheimer’s disease polygenic risk score as a predictor of conversion from mild-cognitive impairment. Translational Psychiatry 2019; 9(1): 154.

8. Mormino EC, Sperling RA, Holmes AJ, et al. Polygenic risk of Alzheimer disease is associated with early- and late-life processes. 2016; 87(5): 481–8.

9. Yu C, Ryan J, Orchard SG, et al. Validation of newly derived polygenic risk scores for dementia in a prospective study of older individuals. Alzheimer’s & dementia : the journal of the Alzheimer’s Association 2023.

10. Xicota L, Gyorgy B, Grenier-Boley B, et al. Association of APOE-Independent Alzheimer Disease Polygenic Risk Score With Brain Amyloid Deposition in Asymptomatic Older Adults. Neurology 2022; 99(5): e462–e75.

11. Kurniansyah N, Goodman MO, Khan AT, et al. Evaluating the use of blood pressure polygenic risk scores across race/ethnic background groups. Nature Communications 2023; 14(1): 3202.

12. Kachuri L, Chatterjee N, Hirbo J, et al. Principles and methods for transferring polygenic risk scores across global populations. Nature reviews Genetics 2024; 25(1): 8–25.

13. Martin AR, Kanai M, Kamatani Y, Okada Y, Neale BM, Daly MJ. Clinical use of current polygenic risk scores may exacerbate health disparities. Nat Genet 2019; 51(4): 584–91.

14. Cosentino S, Schupf N, Christensen K, Andersen SL, Newman A, Mayeux R. Reduced prevalence of cognitive impairment in families with exceptional longevity. JAMA neurology 2013; 70(7): 867–74.

15. Andersen SL. Centenarians as Models of Resistance and Resilience to Alzheimer’s Disease and Related Dementias. Advances in geriatric medicine and research 2020; 2(3).

16. Gunn S, Wainberg M, Song Z, et al. Distribution of 54 polygenic risk scores for common diseases in long lived individuals and their offspring. GeroScience 2022; 44(2): 719–29.

17. Tesi N, van der Lee S, Hulsman M, et al. Cognitively healthy centenarians are genetically protected against Alzheimer’s disease. n/a(n/a).

18. Torres GG, Dose J, Hasenbein TP, et al. Long-Lived Individuals Show a Lower Burden of Variants Predisposing to Age-Related Diseases and a Higher Polygenic Longevity Score. International journal of molecular sciences 2022; 23(18).

19. Wojczynski MK, Jiuan Lin S, Sebastiani P, et al. NIA Long Life Family Study: Objectives, Design, and Heritability of Cross-Sectional and Longitudinal Phenotypes. The journals of gerontology Series A, Biological sciences and medical sciences 2022; 77(4): 717–27.

20. Xicota L, Cosentino S, Vardarajan B, et al. Whole genome-wide sequence analysis of long-lived families (Long-Life Family Study) identifies MTUS2 gene associated with late-onset Alzheimer’s disease. Alzheimer’s & dementia : the journal of the Alzheimer’s Association 2024; 20(4): 2670–9.

21. Daw EW, Anema JA, Schwander K, et al. A Paradigm For Calling Sequence In Families: The Long Life Family Study. bioRxiv 2024: 2024.05.23.595584.

22. Purcell S, Neale B, Todd-Brown K, et al. PLINK: a tool set for whole-genome association and population-based linkage analyses. American journal of human genetics 2007; 81(3): 559–75.

23. Zhou X, Stephens M. Genome-wide efficient mixed-model analysis for association studies. Nature Genetics 2012; 44(7): 821–4.

24. Chen H, Wang C, Conomos Matthew P, et al. Control for Population Structure and Relatedness for Binary Traits in Genetic Association Studies via Logistic Mixed Models. The American Journal of Human Genetics 2016; 98(4): 653–66.

25. Feitosa MF, Kuipers AL, Wojczynski MK, et al. Heterogeneity of the Predictive Polygenic Risk Scores for Coronary Heart Disease Age-at-Onset in Three Different Coronary Heart Disease Family-Based Ascertainments. Circulation Genomic and precision medicine 2021; 14(3): e003201.

26. Ash AS, Kroll-Desrosiers AR, Hoaglin DC, Christensen K, Fang H, Perls TT. Are Members of Long-Lived Families Healthier Than Their Equally Long-Lived Peers? Evidence From the Long Life Family Study. The journals of gerontology Series A, Biological sciences and medical sciences 2015; 70(8): 971–6.

27. Tesi N, Hulsman M, van der Lee SJ, et al. The Effect of Alzheimer’s Disease-Associated Genetic Variants on Longevity. Frontiers in genetics 2021; 12: 748781.

28. Stevenson-Hoare J, Heslegrave A, Leonenko G, et al. Plasma biomarkers and genetics in the diagnosis and prediction of Alzheimer’s disease. Brain : a journal of neurology 2023; 146(2): 690–9.

29. Lumsden AL, Mulugeta A, Zhou A, Hyppönen E. Apolipoprotein E (<em>APOE</em>) genotype-associated disease risks: a phenome-wide, registry-based, case-control study utilising the UK Biobank. eBioMedicine 2020; 59.

30. Sebastiani P, Gurinovich A, Nygaard M, et al. APOE Alleles and Extreme Human Longevity. The journals of gerontology Series A, Biological sciences and medical sciences 2019; 74(1): 44–51.

31. Schächter F, Faure-Delanef L, Guénot F, et al. Genetic associations with human longevity at the APOE and ACE loci. Nat Genet 1994; 6(1): 29–32.

32. Sobel E, Louhija J, Sulkava R, et al. Lack of association of apolipoprotein E allele 4 with late‐onset Alzheimer’s disease among Finnish centenarians. 1995; 45(5): 903–7.

33. Shinohara M, Kanekiyo T, Tachibana M, et al. APOE2 is associated with longevity independent of Alzheimer’s disease. Elife 2020; 9.

34. Sweigart B, Andersen SL, Gurinovich A, et al. APOE E2/E2 Is Associated with Slower Rate of Cognitive Decline with Age. Journal of Alzheimer’s disease : JAD 2021; 83(2): 853–60.

35. Kolbe D, da Silva NA, Dose J, et al. Current allele distribution of the human longevity gene APOE in Europe can mainly be explained by ancient admixture. Aging Cell 2023; 22(5): e13819.

36. Bae H, Song Z, Ali A, et al. Increased genetic protection against Alzheimer’s disease in centenarians. GeroScience 2025.

