## Supplementary Materials for "Utility of Polygenic Risk Scores in Families with Exceptional Longevity"

### Supplementary Table 1. PRS association with Alzheimer disease biomarkers

| | **Log10($A\beta_{40}$)** | | **Log10($A\beta_{42}$)** | | **$A\beta_{42}/A\beta_{40}$** | | **Log10(NfL)** | | **Log(GFAP)** | |
|---|---|---|---|---|---|---|---|---|---|---|
| | Beta | p | Beta | p | Beta | p | Beta | p | Beta | p |
| Kunkle PRS* | 0.000 | 0.843 | -0.001 | 0.510 | 0.000 | 0.291 | 0.001 | 0.734 | 0.003 | 0.148 |
| Kunkle PRS* ≥65 | 0.000 | 0.944 | -0.003 | 0.311 | 0.000 | 0.185 | -0.001 | 0.712 | 0.002 | 0.404 |
| Kunkle Binary | -0.001 | 0.910 | -0.013 | 0.291 | -0.001 | 0.149 | 0.008 | 0.444 | 0.007 | 0.466 |
| Kunkle Binary ≥65 | 0.011 | 0.692 | -0.009 | 0.544 | -0.002 | 0.118 | 0.010 | 0.496 | -0.002 | 0.883 |
| Kunkle 10% | 0.000 | 0.988 | -0.025 | 0.179 | *-0.003* | *0.073* | 0.014 | 0.392 | 0.017 | 0.271 |
| Kunkle 10% ≥65 | -0.010 | 0.684 | *-0.040* | *0.098* | -0.003 | 0.104 | -0.001 | 0.999 | 0.006 | 0.781 |

**Supplementary Table 2: Individual SNP association with Alzheimer's disease biomarkers.**

| Chr | Position | Gene | Kunkle | | LLFS | | | | | | | | | | | |
|---|---|---|---|---|---|---|---|---|---|---|---|---|---|---|---|---|
| | | | | | MAF | OR | AB40 | | AB42 | | AB42/AB40 | | NfL | | GFAP | |
| | | | MAF | OR | | | BETA | PVAL | BETA | PVAL | BETA | PVAL | BETA | PVAL | BETA | PVAL |
| 1 | 207,629,207 | CR1 | 0.187 | 1.17 | 0.177 | 0.94 | -0.006 | 0.506 | *-0.015* | *0.094* | -1.16E-03 | 0.12 | 8.43E-04 | 0.911 | 0 | 0.955 |
| 2 | 127,135,234 | BIN1 | 0.407 | 1.2 | 0.399 | *1.16* | 0.004 | 0.557 | 0 | 0.951 | -3.74E-04 | 0.525 | 6.51E-05 | 0.991 | 0.007 | 0.237 |
| 2 | 233,117,202 | INPP5D | 0.223 | 0.91 | 0.239 | 0.88 | -0.001 | 0.939 | 0.001 | 0.936 | 4.73E-04 | 0.485 | -1.93E-03 | 0.779 | -0.005 | 0.441 |
| 5 | 29,005,878 | SUCLG2P4 | 0.01 | 0.71 | 0.019 | 1 | *0.047* | *0.088* | 0.02 | 0.433 | **-4.79E-03** | **0.034** | -3.12E-02 | 0.171 | -0.017 | 0.431 |
| 5 | 53,369,400 | FST | 0.013 | 0.68 | 0.019 | 0.89 | -0.022 | 0.405 | -0.014 | 0.574 | 8.65E-04 | 0.68 | -6.00E-03 | 0.777 | -0.018 | 0.361 |
| 5 | 88,927,603 | MEF2C | 0.39 | 0.94 | 0.412 | 0.98 | -0.001 | 0.924 | -0.003 | 0.612 | 8.93E-05 | 0.88 | -5.36E-03 | 0.369 | 0.005 | 0.358 |
| 6 | 32,607,629 | HLA -DRB1 | 0.27 | 1.1 | 0.27 | 1.04 | -0.004 | 0.618 | *-0.014* | *0.091* | -1.02E-03 | 0.14 | -3.19E-03 | 0.655 | 0.002 | 0.793 |
| 6 | 41,066,261 | OARD1 | 0.03 | 1.32 | 0.022 | 0.85 | 0.005 | 0.82 | -0.017 | 0.456 | **-3.97E-03** | **0.042** | -7.98E-03 | 0.686 | -0.005 | 0.798 |
| 6 | 41,161,514 | TREM2 | 0.008 | 2.08 | 0.004 | **3.77** | 0.068 | 0.252 | 0.028 | 0.616 | -5.43E-03 | 0.252 | -1.33E-02 | 0.784 | -0.053 | 0.248 |
| 6 | 47,463,548 | CD2AP | 0.28 | 1.09 | NA | NA | NA | NA | NA | NA | NA | NA | NA | NA | NA | NA |
| 7 | 37,804,661 | NME8 | 0.356 | 0.94 | 0.354 | *0.85* | -0.001 | 0.902 | 0 | 0.966 | -2.57E-04 | 0.673 | 8.48E-03 | 0.171 | 0.003 | 0.661 |
| 7 | 100,494,172 | NYAP1 | 0.303 | 0.92 | 0.279 | 1.13 | **0.023** | **0.005** | **0.024** | **0.001** | -3.72E-04 | 0.564 | -6.03E-03 | 0.357 | -0.009 | 0.142 |
| 7 | 143,402,040 | EPHA1 | 0.199 | 0.9 | 0.183 | 1.02 | 0.004 | 0.67 | -0.002 | 0.836 | -5.86E-04 | 0.44 | 7.24E-03 | 0.344 | 0.007 | 0.326 |
| 8 | 27,362,470 | PTK2B | 0.367 | 1.1 | 0.372 | 0.93 | 0.008 | 0.315 | 0.011 | 0.105 | 1.93E-04 | 0.751 | 1.26E-03 | 0.839 | -0.001 | 0.902 |
| 8 | 27,610,169 | CLU | 0.387 | 0.88 | 0.407 | **0.81** | -0.011 | 0.135 | -0.01 | 0.162 | 5.94E-04 | 0.316 | 4.69E-03 | 0.436 | -0.007 | 0.25 |
| 8 | 94,964,023 | NDUFAF6 | 0.476 | 0.94 | 0.45 | 1.05 | -0.008 | 0.268 | -0.011 | 0.101 | -2.01E-04 | 0.73 | -4.03E-03 | 0.5 | -0.001 | 0.908 |
| 10 | 11,678,309 | ECHDC3 | 0.389 | 1.08 | 0.371 | 1 | 0.007 | 0.324 | 0.004 | 0.542 | -3.56E-04 | 0.559 | -2.91E-03 | 0.636 | 0.006 | 0.305 |
| 11 | 47,358,789 | SPI1 | 0.448 | 0.92 | 0.449 | 1.05 | -0.003 | 0.677 | -0.003 | 0.645 | -3.37E-04 | 0.562 | 2.42E-03 | 0.686 | -0.004 | 0.44 |
| 11 | 60,169,453 | MS4A2 | 0.391 | 0.89 | 0.416 | 1.09 | -0.007 | 0.307 | -0.008 | 0.22 | 2.03E-04 | 0.728 | 4.80E-03 | 0.423 | 0.007 | 0.249 |
| 11 | 86,157,598 | PICALM | 0.356 | 0.88 | 0.354 | *0.87* | -0.005 | 0.472 | -0.004 | 0.537 | 1.10E-04 | 0.854 | 4.72E-04 | 0.938 | -0.005 | 0.346 |
| 11 | 121,564,878 | SORL1 | 0.04 | 0.8 | 0.046 | 1.13 | *0.032* | *0.079* | 0.023 | 0.171 | -1.70E-03 | 0.245 | 5.27E-03 | 0.722 | 0.001 | 0.924 |
| 12 | 43,573,874 | ADAMTS20 | 0.413 | 1.06 | 0.41 | 0.89 | -0.01 | 0.172 | -0.004 | 0.535 | 5.48E-04 | 0.354 | 7.37E-03 | 0.219 | -0.001 | 0.88 |
| 14 | 52,924,962 | FERMT2 | 0.093 | 1.14 | 0.098 | **1.38** | 0 | 0.988 | -0.004 | 0.741 | -3.81E-04 | 0.716 | -4.95E-03 | 0.64 | -0.013 | 0.19 |
| 14 | 92,466,484 | SLC24A4 | 0.221 | 0.92 | 0.226 | 0.95 | -0.011 | 0.206 | -0.011 | 0.2 | 3.20E-04 | 0.655 | -1.76E-03 | 0.807 | -0.005 | 0.493 |
| 15 | 50,716,490 | SPPL2A | 0.333 | 0.94 | 0.352 | 1.01 | 0.007 | 0.357 | 0.003 | 0.623 | -9.54E-05 | 0.874 | -3.26E-03 | 0.595 | -0.005 | 0.416 |
| 15 | 58,753,575 | ADAM10 | 0.295 | 0.93 | 0.283 | 0.94 | -0.007 | 0.379 | -0.001 | 0.873 | 8.44E-04 | 0.189 | **-1.35E-02** | **0.039** | -0.006 | 0.316 |
| 16 | 19,796,841 | IQCK | 0.18 | 0.92 | NA | NA | NA | NA | NA | NA | NA | NA | NA | NA | NA | NA |
| 16 | 79,321,960 | WWOX | 0.116 | 1.16 | 0.112 | 0.96 | -0.005 | 0.627 | -0.005 | 0.658 | 7.70E-04 | 0.4 | 1.29E-02 | 0.162 | -0.003 | 0.761 |
| 17 | 58,331,728 | MIR142/TSPOAP1-AS1 | 0.44 | 0.94 | 0.47 | 0.98 | 0.003 | 0.643 | 0 | 0.942 | -7.45E-04 | 0.193 | -1.93E-03 | 0.74 | 0.004 | 0.419 |
| 17 | 63,460,787 | ACE | 0.02 | 1.32 | 0.013 | 0.7 | *0.053* | *0.096* | 0.046 | 0.119 | -2.38E-03 | 0.352 | 2.09E-03 | 0.936 | -0.004 | 0.869 |

| | | | | | | | | | | | | | | | | |
|---|---|---|---|---|---|---|---|---|---|---|---|---|---|---|---|---|
| 19 | 1,056,493 | ABCA7 | 0.182 | 1.15 | 0.173 | 0.95 | 0.005 | 0.623 | 0.006 | 0.492 | 9.61E-04 | 0.218 | 5.90E-04 | 0.941 | 0.001 | 0.878 |
| 20 | 56,422,512 | CASS4 | 0.088 | 0.88 | 0.111 | **0.73** | 0.019 | 0.112 | 0.018 | 0.111 | -1.77E-04 | 0.852 | 7.43E-04 | 0.938 | 0.011 | 0.233 |
| 21 | 26,784,537 | ADAMTS1 | 0.308 | 0.93 | 0.307 | *1.15* | 0 | 0.968 | 0.008 | 0.259 | **1.38E-03** | **0.028** | -2.51E-04 | 0.969 | 0.007 | 0.235 |
